# Left anterior fascicular block is associated with non-ischemic myocardial scar and proportionately decreased ejection fraction

**DOI:** 10.1101/2022.08.02.22277879

**Authors:** Johan von Scheele, Brett D Atwater, Igor Klem, Henrik Engblom, Daniel E Loewenstein, Björn Wieslander, Martin Ugander

## Abstract

**Background:** Left anterior fascicular block (LAFB) has been associated with increased mortality, but the underlying causes are unknown.

**Objectives:** To determine whether LAFB is associated with increased left ventricular (LV) scar burden and reduced LV ejection fraction (LVEF).

**Methods:** LAFB patients (n=51) and matched control patients (n=600) were retrospectively enrolled. Both groups had been referred for cardiovascular magnetic resonance imaging (CMR) and electrocardiography (ECG). They were compared regarding size and location of LV scar, LVEF, and a dysfunction index describing the difference between measured LVEF and expected LVEF based on scar size.

**Results:** Patients with LAFB had on average a larger LV scar (median [interquartile range] 0.7 [0.0-6.6] vs 0.0 [0.0-1.5] % LV mass, p<0.001). LAFB was associated with a higher prevalence of any scar (59% vs 33%, p<0.001). The groups had similar prevalence of ischemic scar (29% vs 23%, p=0.40) but LAFB patients a higher prevalence of non-ischemic scar (29% vs 10%, p=0.001) most frequently located in the basal and mid inferoseptal segments and the anterior and lateral apical LV segments. LVEF was lower in LAFB than in controls (58 [43-60] vs 60 [55-60] %, p=0.02). There was no difference in dysfunction index (24.0 [17.8-25.5] vs 24.0 [19.0-27.8] %-points of LVEF, p=0.32).

**Conclusions:** In a matched cohort, LAFB was associated with a small decrease in LVEF that was proportionate to the increased LV scar burden, which was more commonly of non-ischemic etiology and not infarction, and not more commonly located near the expected course of the left anterior fascicle.

## Introduction

Left anterior fascicular block (LAFB) is a cardiac conduction abnormality with slow or absent conduction in the anterior fascicle of the left bundle branch, resulting in delayed activation of the left ventricular (LV) anterior or anterolateral wall, shifting the electrical axis in the frontal plane leftward. This conduction block is diagnosed by electrocardiography (ECG) as a frontal plane QRS axis between -45 and -90 degrees, and other associated ECG markers. LAFB is a relatively common conduction abnormality and its prevalence increases with age^1^. LAFB has been reported in 4-6% of the general population aged 60 years or older, and in 10% of people aged 80 years or older^2^. Unlike complete left bundle branch block, LAFB has long been regarded as a benign ECG finding that has a limited association with increased morbidity or mortality when not combined with other risk factors^3-5^. However, in recent years this view has been challenged. LAFB has relatively recently been shown to be related to all-cause and cardiovascular mortality as well as incident atrial fibrillation^6^. The underlying pathophysiological mechanisms by which LAFB is associated with these outcome measures are still not known. It has previously been indicated that LAFB might be associated with increased fibrosis in the LV myocardium at histopathology^7,8^. However, histopathological studies have been limited in cohort size and have not found any clearly focal distribution of the fibrosis near the course of the anterior fascicle associated with LAFB^7-9^.

Currently, the use of cardiac magnetic resonance imaging (CMR) with late gadolinium enhancement (LGE) enables non-invasive detection of focal myocardial fibrosis of both ischemic and non-ischemic etiology^10,11^.

The aim of this study was to address the hypothesis that LAFB is associated with increased LV scarring and decreased LV ejection fraction (LVEF) compared to matched control patients, and to investigate whether LAFB is associated with a particular type of scar pattern or myocardial location of scar.

## Methods

### Study cohort and design

In this retrospective study, n=651 patients were retrospectively and consecutively included from a clinical database of patients undergoing CMR at Duke University Medical Center (NC, USA). All patients had undergone resting ECG between 2012 and 2014. All patients (51 patients with LAFB and 600 matched controls without LAFB) had a LGE CMR examination performed within -1 to 180 days of the ECG recording. LAFB was defined by 12-lead ECG as a frontal plane QRS axis between -45 to -90 degrees in the absence of LV hypertrophy on ECG, and with QRS duration less than 120 ms. Exclusion criteria for the control group were: LAFB, left bundle branch block (LBBB), right bundle branch block (RBBB), RBBB+LAFB, left posterior fascicular block, or pre-excitation on the ECG recorded closest to the CMR examination. Furthermore, controls were statistically matched to the LAFB group regarding age and sex using an exact matching method. Further exclusion criteria included subjects with asymmetric or apical LV hypertrophy on CMR. All CMR images were reported by a board-certified cardiologist with level 3 CMR training. The study was approved by the Institutional Review Board at Duke University, Durham, North Carolina, USA (IRB# 48875), with a retrospective waiver of individual informed consent.

### Scar assessment

The presence and location of hyperenhanced tissue on LGE, which was interpreted as representing scarred myocardium, was determined by visual inspection using the AHA 17-segment model^12^. Regional LGE was scored according to the spatial extent within each segment (0=no hyperenhancement, 1=1-25% hyperenhanced, 2=25-50%, 3=50-75%, 4=75-100%). Scar size as a percentage of LV myocardium was then calculated by summing the segments with hyperenhancement (each weighted by the midpoint of the range of hyperenhancement for the given segmental score, i.e. 1=12.5%; 2=37.5%; 3=62.5%; 4=87.5%) and dividing by 17^13^.

### LVEF and dysfunction index

LVEF was assessed visually after assessment of cine images in multiple projections. A dysfunction index was defined as the difference between expected maximum LVEF and estimated LVEF as previously described^14^. Briefly, expected maximum LVEF was determined iteratively by the straight line on a graph of scar versus LVEF yielding the smallest area under the line while simultaneously satisfying the condition of letting at least 95% of all control patients with >0% scar be found under the line (Figure 1). A larger dysfunction index is seen when factors other than scar play a larger role in decreasing the LVEF. One possible factor contributing to an increased dysfunction index is dyssynchronous contraction of the myocardium causing a decreased pumping efficiency. Consequently, when the dysfunction index is low, there is a low likelihood that factors other than scar play a role in reducing LVEF.

**Figure 1.**
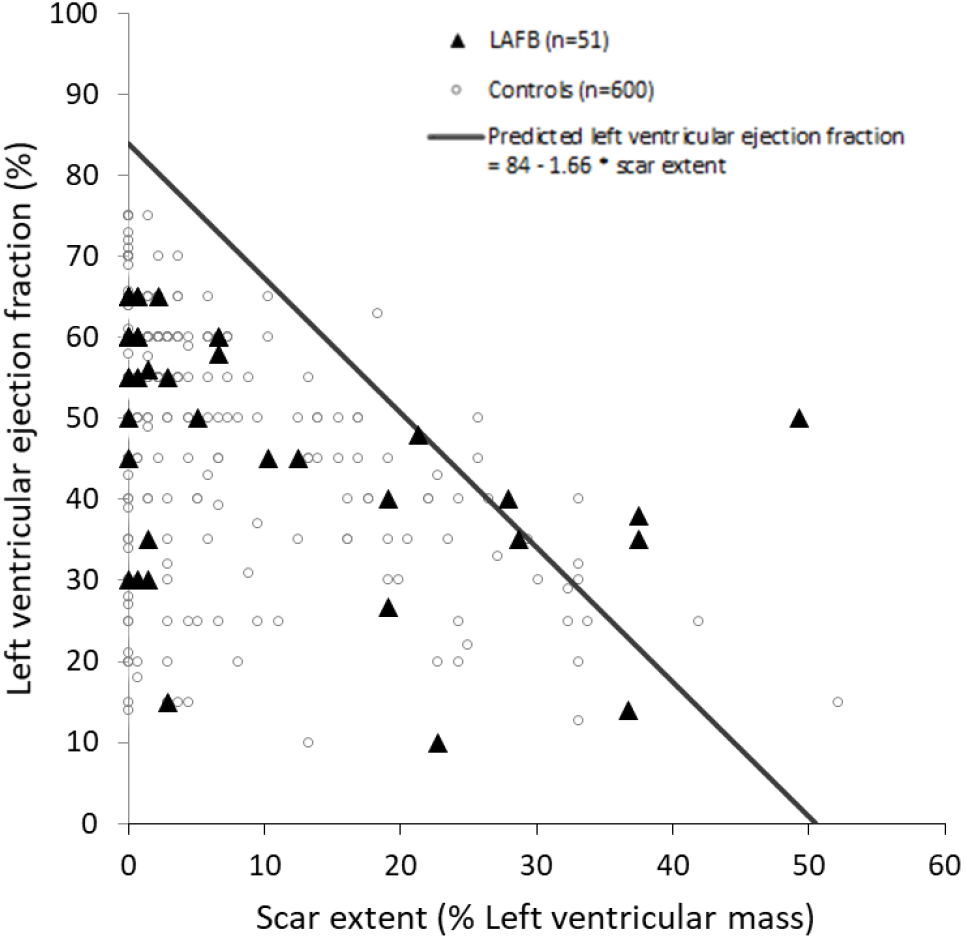
Relationship between ejection fraction and scar for left anterior fascicular block (LAFB) and controls. The diagonal grey line represents the maximum predicted ejection fraction in relation to scar extent for the control population. See methods for details of how this line was determined. Open circles denote data points for controls, and black triangles denote data points for patients with LAFB. Note, multiple data points may be identical and overlapping.

### Statistical analysis

Statistical analyses were performed in the software R (RStudio 1.2.5042, Boston, MA, USA). The Shapiro-Wilk test was used for assessing normality of distribution. Non-normally distributed continuous measures are reported as median [interquartile range], and differences were tested using the Mann-Whitney U test. Exact matching was performed using the MatchIt package in R^15^. Differences in prevalence were tested using the Chi squared test or Fischer’s exact test, as appropriate. For statistical tests, a p-value <0.05 was considered statistically significant except for segmental scar analysis where Bonferroni correction was used to correct for multiple tests. For these analyses, a p-value <0.05/17 (0.003) was considered statistically significant for segmental scar analysis in the 17-segment model.

## Results

Patient characteristics are presented in Table 1. A total of 1612 subjects underwent matching (n=51 LAFB and n=1561 controls), resulting in a LAFB group (n=51) and a matched control group (n=600). Compared to matched controls, LAFB was associated with a higher prevalence of heart failure (HF). All other evaluated patient characteristics showed no difference in frequency between LAFB and controls.

**Table 1.** Demographics and prevalence of clinical covariates.

| Covariate | LAFB | Controls | p-value |
| --- | --- | --- | --- |
| **Demographics** |  |  |  |
| Number of patients, n | 51 | 600 |  |
| Age, years | 69 [60-73] | 68 [61-71] | 0.54 |
| Male sex, n (%) | 35 (69) | 387 (65) | 0.66 |
| Time from ECG to CMR, days | 4 [0-22] | 7 [0-37] | 0.51 |
| **Past Medical History, n (%)** |  |  |  |
| Atrial flutter or fibrillation | 26 (51) | 324 (54) | 0.79 |
| Chronic obstructive pulmonary disorder | 10 (20) | 142 (24) | 0.63 |
| Coronary artery disease | 35 (69) | 335 (56) | 0.10 |
| Diabetes | 17 (33) | 211 (35) | 0.91 |
| Heart Failure | 29 (57) | 211 (35) | **0.003** |
| Hypertension | 37 (73) | 485 (81) | 0.21 |
| Obstructive sleep apnea | 13 (25) | 189 (32) | 0.46 |
| **Medications, n (%)** |  |  |  |
| ACE Inhibitor | 28 (55) | 252 (42) | 0.10 |
| Amiodarone | 4 (8) | 43 (7) | 0.78 |
| Beta blocker | 38 (75) | 396 (66) | 0.28 |
| Flecainide or propafenone | 3 (6) | 47 (8) | 0.79 |
| Dofetilide or sotalol | 5 (10) | 81 (14) | 0.60 |

Continuous data are presented as median [interquartile range].

### LV scar size and systolic function

The maximum predicted LVEF in relation to scar for the control population was found to be described by the formula LVEF = 84 – 1.66 * scar extent (Figure 1). Compared to controls, patients with LAFB had reduced LVEF (58 [43-60] vs 60 [55-60] %, p=0.02) and an increased amount of LV scar (0.7 [0.0-6.6] vs 0.0 [0.0-1.5] % LV mass, p<0.001). However, there was no difference in dysfunction index between LAFB and controls (24.0 [17.8-25.5] vs 24.0 [19.0-27.8 %-points of LVEF, p=0.32) (Figure 2).

**Figure 2.**
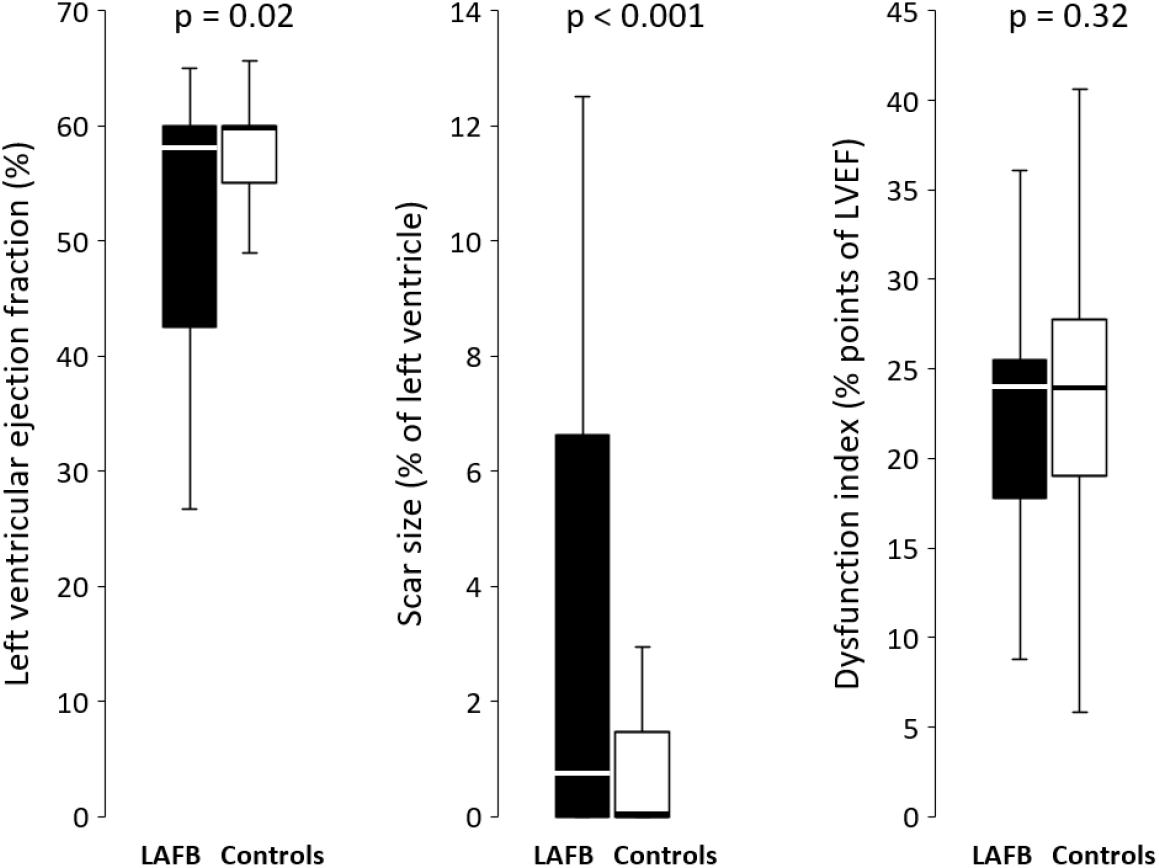
Left ventricular ejection fraction, scar size, and dysfunction index for left anterior fascicular block (LAFB) and controls. Data are shown as box and whisker plots where the box indicates the median and interquartile range, and the whiskers indicate the range.

### LV scar etiology and location

LAFB was associated with a higher prevalence of any scar (59% vs 33%, p<0.001), and this was due to a higher prevalence of non-ischemic scar (29% vs 10%, p=0.001) but not more scar due to myocardial infarction (29% vs 23%, p=0.40) (Figure 3, Central Illustration). Figure 4 shows the comparison of the amount of scarring for each of the 17 segments of the LV. LAFB showed a higher prevalence of scar in the basal inferoseptal, mid inferoseptal, and apical lateral segments (p<0.003 for all). Those same segments, as well as the apical anterior segments, were all associated with a higher prevalence of non-ischemic scar in LAFB (p<0.003 for all). There were no differences in scar prevalence due to myocardial infarction between any of the 17 segments when comparing LAFB to controls (p>0.003 for all).

**Figure 3.**
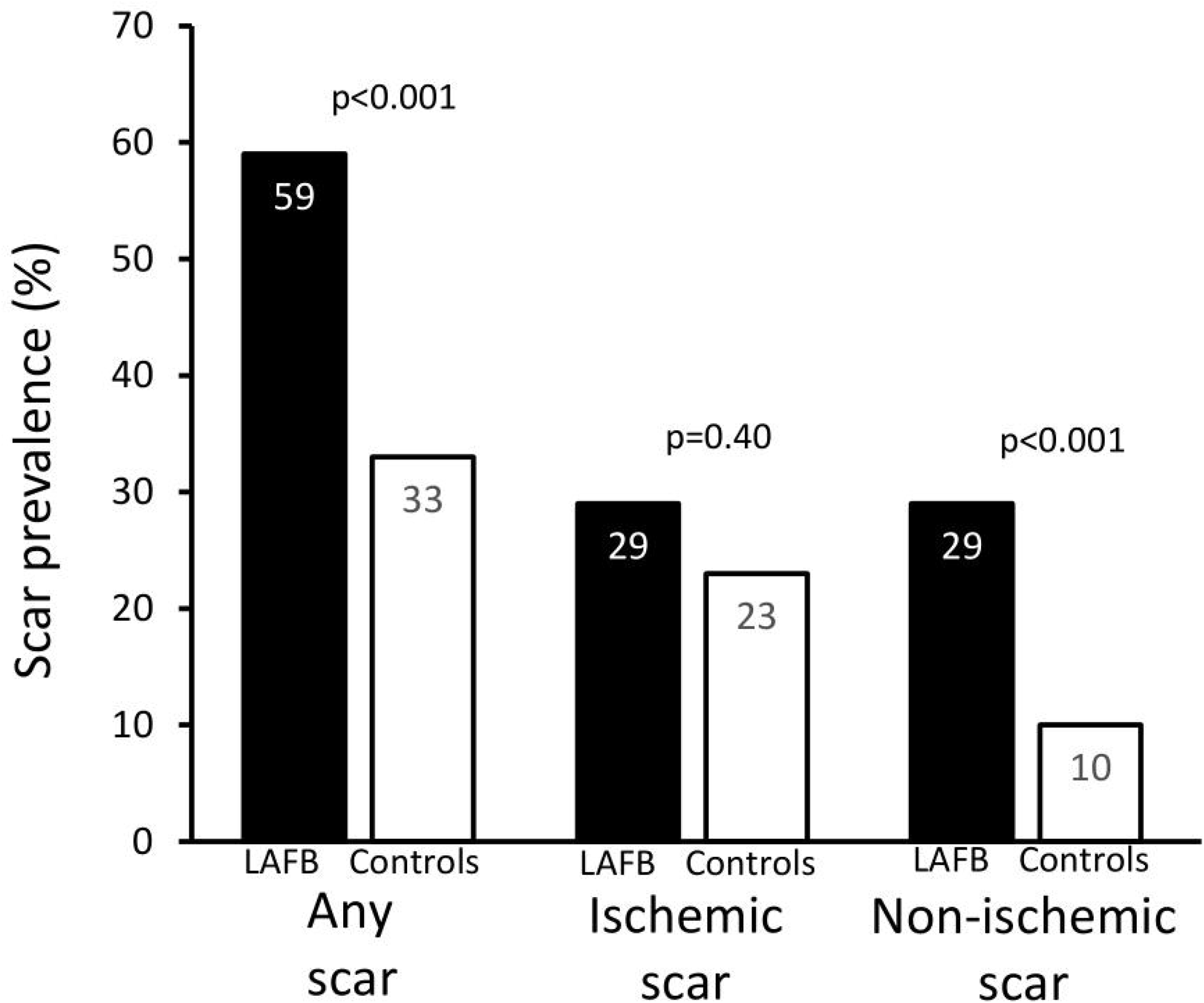
Prevalence of any type of scar, ischemic pattern scar and non-ischemic pattern scar for left anterior fascicular block (LAFB) and controls.

**Figure 4.**
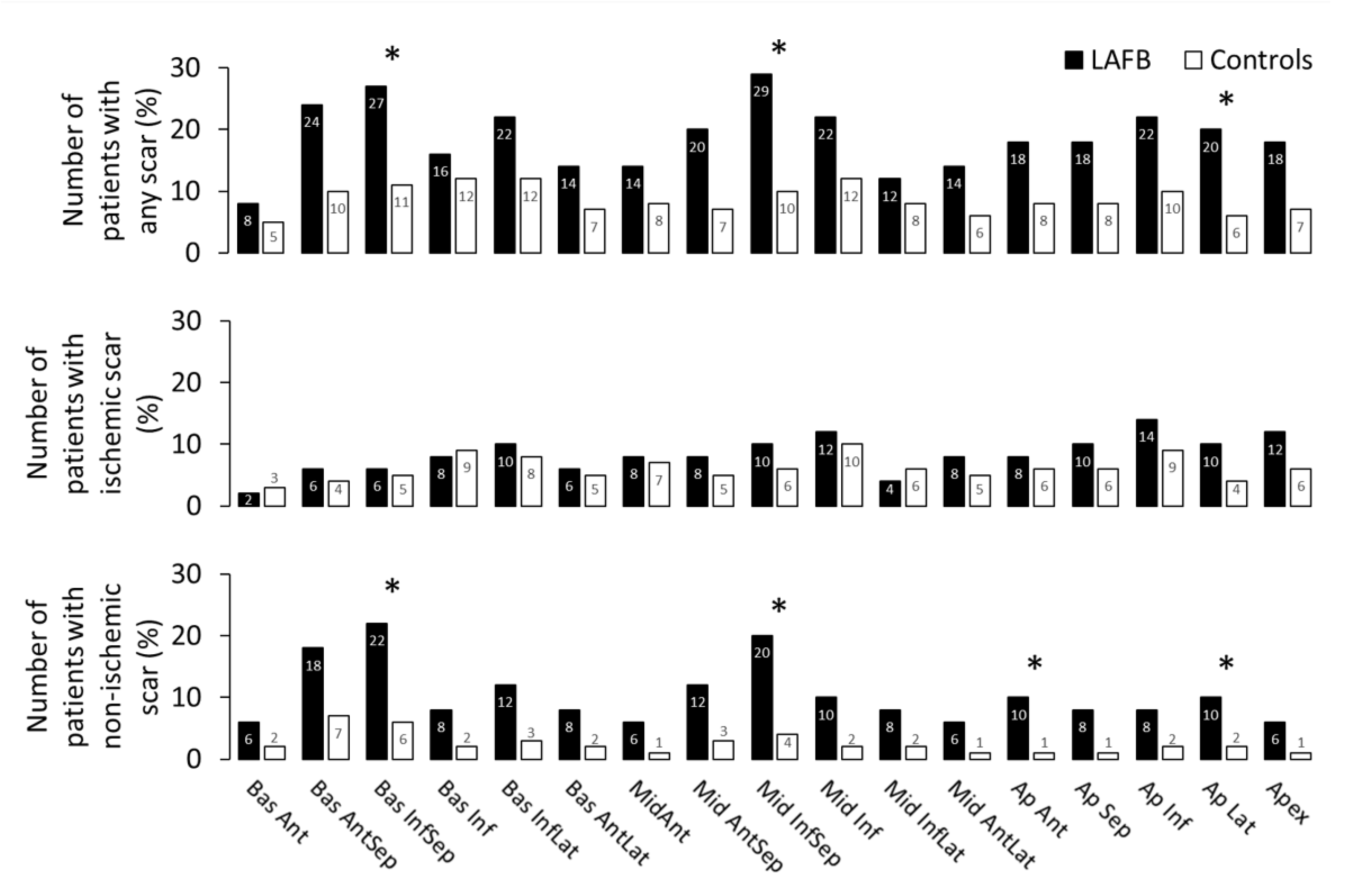
Prevalence of scarring across individual myocardial segments for left anterior fascicular block (LAFB) and controls. Bas = basal, Mid = midventricular, Ap = apical, Ant = anterior, Sep = septal, Inf = inferior, Lat = lateral.

## Discussion

The main finding of the study is that in a hospital-based cohort of patients having undergone CMR, LAFB was associated with a decreased LVEF that was proportionate to the increased LV scar burden, and which was more commonly due to non-ischemic etiology and not infarction, and not more commonly located near the origin of the left anterior fascicle.

### LV scar size, etiology, and location

The current study found that LAFB is associated with increased LV scar size and reduced LVEF. Differences in LV scar size were driven by a higher prevalence of non-ischemic pattern scar. Interestingly, there was no increase in prevalence of scarring in the basal anteroseptal and anterior segments of the LV, which are those segments where the left anterior fascicle has its course^16^. It thus seems unlikely that focal non-ischemic scarring or infarction detectable by LGE CMR along the course of the left anterior fascicle is the primary etiology of LAFB. Instead, LAFB may be an ECG marker of diffuse myocardial fibrosis or functional block of the conduction system seen e.g. in aging or in what sometimes is referred to as Lenègre’s and/or Lev’s disease^17^, and which is not visible by LGE CMR.

### Dysfunction Index

The dysfunction index makes it possible to quantitatively compare the influence of LV scarring on the reduction in LVEF in relation to causes other than scarring. This comparison is performed in order to assess whether scar associates as much to reduced LVEF in LAFB patients as it does in patients with normal conduction.

The dysfunction index has recently been found to be increased in LBBB compared to controls^18^. Those findings imply that factors other than scar (e.g. LV dyssynchrony) play a role in reducing LVEF in LBBB. Prior work shows that more than 10% of the patients with a LVEF <35% and QRS duration <120ms exhibit interventricular dyssynchrony, and nearly 30% have intraventricular dyssynchrony^19^. If LAFB caused LV dyssynchrony, LAFB would have been expected to have an increased gap between measured and expected maximum LVEF, and subsequently an increased dysfunction index. That would indicate that the reduction in LVEF could be influenced by factors other than scar. However, this was not the case in our study. The dysfunction index was similar in patients with LAFB and matched controls suggesting that LAFB does not have an additional effect beyond its association with increased LV scar on lowering global LV function measured as LVEF.

### Clinical covariates

The current study identified a control group that was matched for age and sex. However, the one exception to matching was that LAFB was more often associated with a clinical diagnosis of HF. The increased prevalence of HF in the LAFB group goes hand in hand with the demonstrably lower LVEF and a larger scar size. It is possible that comorbidities or factors other than those available for comparison herein were prevalent to a higher or lower degree among either LAFB or controls.

### Limitations

The findings in the present study should be interpreted in the light of some limitations. First, the 180-day period allowed between ECG and CMR is a limitation of the study as the electrical axis may change or other diagnoses arise on ECG over time. Ideally, all patients would have had ECG performed the same day as CMR. However, the median difference in time between ECG and CMR was low for both groups (four and seven days, respectively). This suggests that time difference between CMR and ECG was not a major factor.

Another factor that may influence the results is the fact that all patients were recruited from a hospital population having undergone both ECG and CMR with LGE. This could mean that both study groups on average are less healthy than a primary care outpatient population or the general population, in which case the study may be less generally applicable.

The study was limited by a relatively modest sample size in the LAFB group. However, LAFB represented 3% of the whole cohort before the matching process, which is in line with what could be expected from the prevalence in the general population^1^, and no LAFB subjects were excluded in the matching process.

Despite matching for age and sex, patients with LAFB had a higher prevalence of HF than matched patients with normal conduction. This may explain the higher frequency of LV scar and the lower LVEF observed in the LAFB cohort.

## Conclusions

In conclusion, in a hospital cohort referred for CMR, LAFB is associated with an increased scar burden and a proportionately reduced LVEF. LV scarring was more often present in a non-ischemic pattern. There was not an increased presence of scar in the location of the left anterior fascicle, demonstrating that macroscopic focal scarring detectable by LGE CMR is not a common cause of LAFB.

## Clinical competencies

This study of left anterior fascicular block (LAFB) used cardiovascular magnetic resonance imaging to demonstrate that LAFB is not associated with an increased amount of myocardial infarction, but is associated with an increased prevalence of focal myocardial scarring of non-ischemic origin. However, this scarring was not more commonly located near the left anterior fascicle, and thus could not be causally linked to LAFB. ACGME clinical competency: medical knowledge.

## Translational outlook

This study provides insight into the etiologies associated with LAFB. As LAFB is a frequently encountered conduction disturbance, it is important to understand the underlying etiology in order to inform further clinical management. It may be that a more nuanced understanding of LAFB is required in order to more effectively determine whether there exists a sub-group of patients at risk within the wider population, and future studies to address this are warranted.

## Data Availability

Data produced in the present study may be available upon reasonable request to the authors if permissible.

## Abbreviations

LAFB: Left anterior fascicular block
LV: Left ventricle
ECG: Electrocardiography
CMR: Cardiovascular magnetic resonance imaging
LGE: Late gadolinium enhancement
LVEF: Left ventricular ejection fraction
LBBB: Left bundle branch block
RBBB: Right bundle branch block
HF: Heart failure

## Figure legends

**Table 1. Patient demographics and prevalence of clinical covariates**. Continuous data are presented as median [interquartile range].

**Table 1 abbreviations:** ACE – Angiotensin-converting-enzyme, CMR – Cardiovascular magnetic resonance imaging, ECG – Electrocardiography, LAFB – Left anterior fascicular block

## Central Illustration

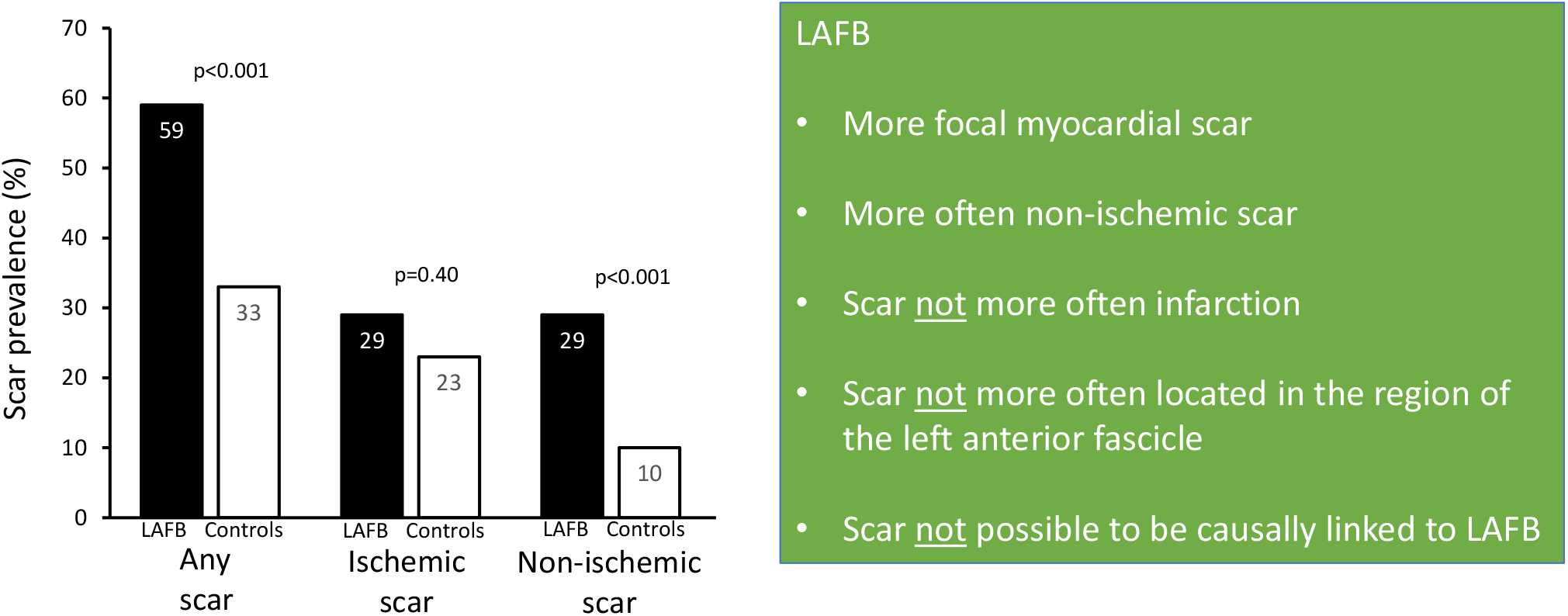

## Notes

### Competing Interest Statement

Dr. Ugander has a development agreement for CMR between Karolinska University Hospital and Siemens Healthineers. Dr. Atwater is an advisory board member at Medtronic, Biotronik, Biosense Webster and Abbott and a consultant at Abbott. The remaining authors have no relationships to declare.

### Funding Statement

This study did not receive any external funding.

### Author Declarations

IRB of Duke University, Durham, North Carolina, USA, gave ethical approval for this work.

### Summary of Updates

Mainly formatting. No new data, no new scientific content. Updated conflicts of interest.

